# Domain-wide Mapping of Peer-reviewed Literature for Genetic Developmental Disorders using Machine Learning and Gene2Phenotype

**DOI:** 10.1101/2025.11.24.25340871

**Authors:** T. Michael Yates, Sarah E. Hunt, Diana Lemos, Seeta Ramaraju Pericherla, Elena Cibrian Uhalte, Morad Ansari, Louise Thompson, Simona E. Doneva, Caroline F. Wright, Helen V. Firth, T. Ian Simpson

## Abstract

Genetically determined developmental disorders (GDD) are rare, heterogeneous conditions for which clinical diagnosis increasingly depends on genomic variant prioritisation and rapid synthesis of genotype–phenotype evidence scattered across the literature. Manual curation of this evidence is labour-intensive, difficult to scale and to keep up to date. We present a domain-wide, automated pipeline that identifies PubMed abstracts describing human case reports/series and maps them to molecularly defined diseases in the Gene2Phenotype database (G2P). The natural language processing system combines a fine-tuned BERT classifier (LitDD BERT) to detect GDD-relevant abstracts, a fine-tuned cross-encoder (LitDD Crossencoder) to propose disease candidates, and a constrained large language model to adjudicate final mappings. Trained on 13,738 annotated title–abstract pairs spanning 231 genes and 354 G2P entries, LitDD BERT achieved an F1 of 0.89 (precision 0.83, recall 0.94). The cross-encoder reached 0.99 top-5 recall, and the full ensemble attained precision 0.89, recall 0.82, and F1 0.85 on a held-out test set. Applied to the entirety of PubMed, the pipeline identified approximately 69,000 manuscripts which could be mapped to G2P diseases. Against independent manually curated sets, it retrieved about 70% of manuscripts at both micro and macro levels, indicating generalization across diverse disease mechanisms, differing curation standards and inheritance patterns. The resulting corpus is accessible through G2P and is being integrated into routine biocuration. This work delivers scalable, updateable literature surveillance for GDD, enabling faster evidence review in a diagnostic setting, supporting bioinformatic pipelines, and laying a foundation for downstream case-level extraction and standardized phenotype integration.

## Introduction

Diagnosis in genetic disease relies on clinical interpretation of an individual’s phenotypic match to genomic variants identified. Given that each genome contains 4-5 million variants (1), genomic diagnostic sequencing has necessitated the development of complex bioinformatic filtering pipelines to reduce candidate disease variants to a manageable number (2,3). However, phenotype analysis still often requires time-consuming manual review of case reports/series in the peer-reviewed literature, to compare a patient’s phenotype to those reported with similar variants, resulting in significant clinical workload (4,5).

Genotype-phenotype databases have been developed to address this issue, defining disease-gene relationships and collating evidence for these assertions, which often takes the form of published phenotypic series (6–8). More recently, an international collaboration, the Gene Curation Coalition, has been established to harmonize these curation efforts (9). This synthesis and systematic analysis of evidence reduces clinician time spent searching the primary literature, as well as allowing for incorporation of disease/phenotype data into bioinformatic pipelines. However, creating and updating these gene-disease association databases is heavily reliant on manual biocuration (8).

The Gene2Phenotype (G2P) database (https://www.ebi.ac.uk/gene2phenotype) was established over a decade ago to compile and share monogenic gene-disease associations, including phenotype and detailed molecular mechanistic data (8). These assertions are categorised into disease domains and evidenced by links to the primary literature using PubMed IDs (PMIDs) (10). Literature search is currently a manual process, which is highly resource intensive. This is particularly true for updating existing entries which have limited confidence and do not meet the threshold for clinical reporting. This curation process is especially challenging in the domain of genetically-determined developmental disorders (GDD). These are rare conditions which arise during pregnancy or early life. GDD are highly genotypically and phenotypically heterogeneous, with approximately 2800 conditions currently described in Developmental Disorders G2P (DDG2P) (8). Newly defined GDD continue to be described through case series/case reports in the peer-reviewed literature, in addition to new reports of known diseases. G2P is updated twice a month with the latest curations.

Natural language processing, a subfield of artificial intelligence, has been increasingly applied to the analysis of medical literature. This includes classifying abstracts for use in systematic reviews (11,12), as well as to aid curation of genetic disease (13–15).

The recent emergence of large language models (LLM) has led to work utilising their deep contextual reasoning abilities to analyse manuscripts relevant to genetic disorders (13). There remain several issues to overcome, for example standardising data despite the heterogeneity of disease descriptions and distinguishing between manuscripts which describe human subjects rather than functional or molecular work. Additionally, the challenge of mapping relevant papers to specific genetic diseases, rather than just genes, remains unsolved. This is a complex topic, with no universally agreed definition of disease or nomenclature (16–18).

Here, we present a PubMed-wide mapping of case reports/case series across the entire spectrum of GDD, using molecularly defined disease definitions from G2P. We utilise a pipeline finetuned on a corpus of manually annotated GDD abstracts. We first train a BERT model to learn the specific language patterns found in abstracts of genetic disease-specific articles and use this to screen the entire medical literature in PubMed (c. 38 million articles) (19). Next, we map these abstracts to DDG2P diseases using a cross-encoder. This compares the textual representation of every condition in DDG2P against the article abstracts to generate a similarity score (20). The highest scoring candidate mappings undergo final arbitration using the DeepSeekR1 large language model (LLM) (21). The pipeline allows for scalable and easily updated literature classification. This should prove useful not only in biocuration of disease databases, but also to clinicians and clinical scientists searching the primary literature, and for use in bioinformatic pipelines/methods.

## Methods

### Data collection and annotation

A corpus of PubMed (10) records was created using a subset of genes from DDG2P (downloaded 4^th^ August 2025). Previous DDG2P curation experience was used to select a set of genes thought to represent both the clinical spectrum of conditions described in the database and disease entities which might be more challenging for curators as well as the classifier. Well established conditions (likely described in multiple papers) were represented by the 50 most common diagnoses in the Deciphering Developmental Disorders (DDD) study (3). Newly described conditions were included (often reported in only a single paper) from DDG2P entries created within the past 2 years. Additionally, diseases which are challenging to curate were selected randomly from entries with multiple allelic requirements for the same phenotype, and genes with pleiotropy/allelic disorders, i.e. multiple diseases per gene.

For each of these genes, a PubMed search for the HGNC gene symbol in the title was performed, using NCBI (National Center for Biotechnology Information) E-utilities (10,22). This search was thought likely to be enriched for GDD case reports/series for a given condition, from previous curation experience. Manuscripts published before 1980 were discarded as these were thought to be more likely to contain phenotypic descriptions without a molecular confirmation of the genetic basis of the disease.

Each gene was mapped to all corresponding DDG2P entries. For each DDG2P disease, the title and abstract (TIAB) of the first 100 PubMed results (or all results if less than 100) were annotated by a Clinical Geneticist experienced in gene-disease curation (T.M.Y.). Annotations were using binary labels (1/0). The positive label (1) was used if the TIAB described case reports for the relevant DDG2P entry or if the title/abstract appeared to include case reports for the relevant DDG2P entry but also included new reports for other diseases, or if the paper was a review. The negative label (0) was if the TIAB did not describe any case report data for the relevant DDG2P entry.

### Model finetuning

This dataset was used to train models for incorporation into a pipeline and to test performance on an unseen set (Figure 1). Each model was selected using baseline performance benchmarking. A label-stratified split was used to create train and test sets of 80%/20%. The same training set was used for BERT and crossencoder models, given these had different tasks (classification/disease mapping). ModernBERT (19) was chosen as the base model for fine-tuning a GDD classifier. Fine tuning of the annotated corpus with a BERT classification head was performed using HuggingFace (23). Hyperparameter optimisation was with Ray Tune (24).

**Figure 1:**
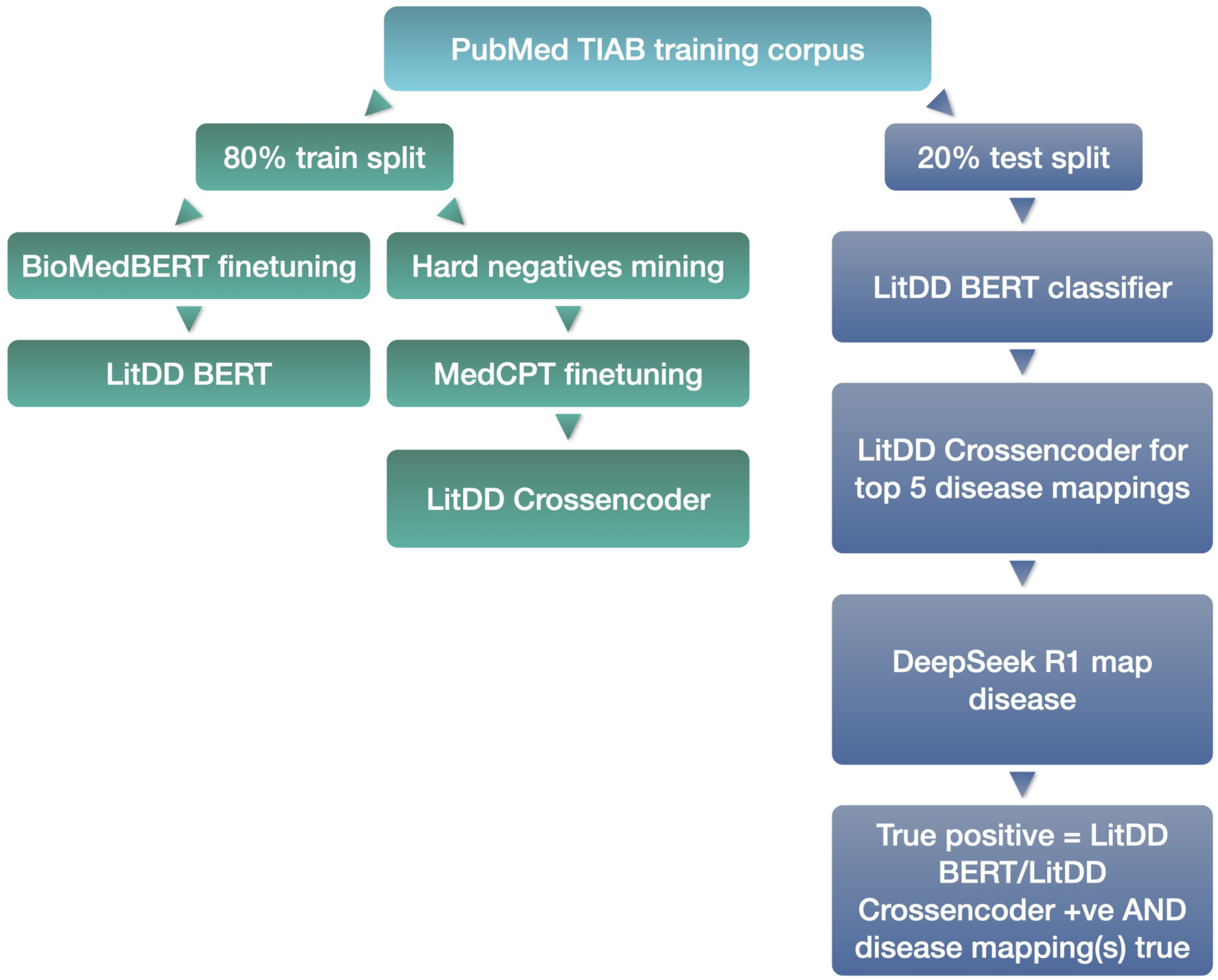
Train and test workflow for identifying PubMed TIAB describing GDD and mapping to DDG2P disorders (8). A corpus of TIAB identified through PubMed gene symbol[title] search were annotated by a clinical geneticist for a subset of DDG2P genes. Annotations were binary - TIAB includes mention of newly-described individuals with a GDD or not. The train set was used to finetune a ModernBERT model (19). The resulting classifier - LitDD BERT - identifies TIAB as describing GDD or not. For the crossencoder, the train dataset was mapped to DDG2P disorders. The data was enhanced using hard negatives mining with a biencoder (25) to enable better differentiation between similar correct/incorrect TIAB-disease pairs. A biomedical crossencoder - MedCPT (20) - was finetuned on this set, generating similarity scores for a TIAB-DDG2P disease input. To test performance on unseen data, LitDD BERT classified abstracts as GDD-relevant or not. LitDD crossencoder identified the top five DDG2P mappings for each TIAB by comparing the text to all disorders in the database. The top five by similarity score were then analysed by the DeepSeek R1 LLM (21) to generate the final mapping(s). For testing purposes, a TIAB was a true positive if LitDD BERT classified positive, the LitDD Crossencoder disease similarity score was greater than 0.9, and the resulting disease mapping(s) matched those from manual annotation. TIAB - title + abstract, GDD - genetically-determined developmental disorders, DDG2P Developmental disorders in G2P database, LLM - large language model.

Each entry in the crossencoder training set was linked to a DDG2P locus-disease-mechanism-genotype-evidence (LGMDE) thread (8) via the unique G2P ID. The LGMDE thread defines a disorder in G2P through inclusion of gene, mechanistic information, phenotype and disease name, using standardised terminology. Hard negative mining was performed on the crossencoder training set using a MedEmbed biencoder model (25). All LGMDE threads in DDG2P were added as a source of additional negatives. Hard negative mining aims to identify anchor-negative pairs - in this case TIAB and LGMDE threads - which are difficult for the model to differentiate. This leads to increased performance in finetuning (26).

The DeepSeek-R1-Distill-Qwen-14B LLM (21) was used directly on the test set to identify the DDG2P disease or diseases described in a TIAB. A custom prompt was used, including the top five crossencoder full LGMDE mappings and the text of a given TIAB.

Performance metrics used true positives defined as BERT positive, crossencoder score >= 0.9 (corresponding to a clear binary split of papers on manual review), and all diseases corresponding to final G2P ID mappings contained within abstract, as defined by manual annotation.

### Mapping PubMed to DDG2P

The models described above were used to create a pipeline for classifying the entirety of PubMed and mapping relevant papers to DDG2P (Figure 2). The LitDD BERT model was used to classify all TIAB in a copy of PubMed MedLine records (downloaded August 2025). For each TIAB classified positive, the LitDD Crossencoder was used to generate similarity scores vs all DDG2P LGMDE (downloaded August 2025). The top five LGMDE by score were then given as input to the LLM which was asked to identify the correct LGMDE thread(s) described by the TIAB. The final mappings used the true positive criteria described above. The data was cleaned by ensuring all G2P ID mappings were true, and the gene corresponding to the mapping was mentioned in the TIAB.

**Figure 2:**
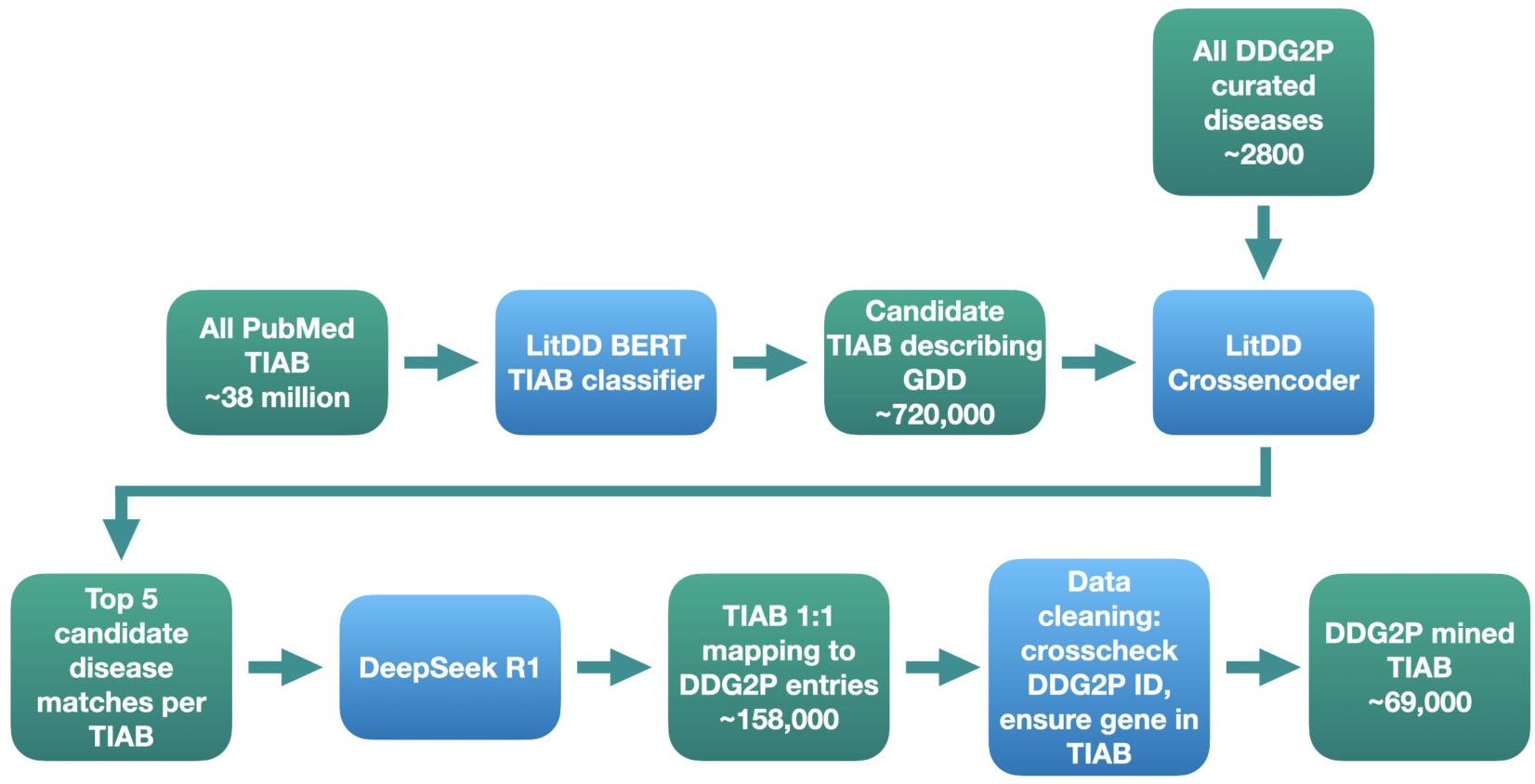
Pipeline for PubMed-wide identification of TIAB describing developmental disorders and mapping to DDG2P diseases (8,10). The finetuned LitDD BERT model was used to classify all PubMed TIAB as containing newly described subjects with a GDD or not. Each classified positive TIAB was then compared to every disorder in DDG2P using similarity scores generated by the finetuned LitDD Crossencoder. The top five TIAB by score were then analysed by DeepSeek R1 (21) to generate a final disease mapping(s). The mappings provided by the LLM were verified against DDG2P. Only TIAB containing mention of the corresponding gene(s) were included, as defined by PubTator (15). This pipeline therefore allows mapping of the entire developmental disorders peer-reviewed literature as defined by DDG2P. TIAB - title + abstract, GDD genetically-determined developmental disorders, DDG2P - Developmental disorders in G2P database, LLM - large language model.

## Results

### Performance of classification and mapping pipeline

The annotated train/test dataset was derived from 231 genes corresponding to 354 DDG2P entries. PubMed gene symbol [title] searches for these genes resulted in 13738 unique annotated TIAB, of which 3534 described a DDG2P disorder (26%).

ModernBERT and NCBI MedCPT (19,20) were chosen as the classification/crossencoder models respectively, based on baseline performance against similar models (Table 1, Table 2). Of note, ModernBERT is not biomedical domain specific, unlike the well-known BioBERT and BioMedBERT (27,28). However, ModernBERT was trained using a much larger dataset than these older models, likely to contain biomedical domain-specific information. Finetuning significantly improved performance (Table 1, Table 2).

**Table 1:** Performance of LitDD classifier against baseline models (19,27,28) on a custom annotated test set. Metrics for LitDD are means over 5-fold cross validation.

| Model | Precision | Recall | F1 |
| --- | --- | --- | --- |
| BioBERT | 0.16 | 0.14 | 0.15 |
| BioMedBERT | 0.09 | 0.20 | 0.12 |
| ModernBERT (baseline) | 0.11 | 0.31 | 0.16 |
| LitDD BERT (finetuned) | **0.83** | **0.94** | **0.89** |

**Table 2:** Performance of LitDD Crossencoder classifier against baseline models (20,29,30) on a custom annotated test set. Top k is by crossencoder score, indicating presence of correct disease mapping(s) in list of k crossencoder candidates. Metrics for LitDD Crossencoder are means over 5-fold cross-validation.

| Model | Top-1 | Top-2 | Top-3 | Top-4 | Top-5 |
| --- | --- | --- | --- | --- | --- |
| MediMaven ReRanker | 0.14 | 0.20 | 0.23 | 0.25 | 0.27 |
| MS Marco MiniLM | 0.21 | 0.27 | 0.29 | 0.30 | 0.31 |
| NCBI MedCPT (baseline) | 0.69 | 0.89 | 0.95 | 0.97 | 0.98 |
| LitDD Crossencoder (finetuned) | **0.76** | **0.93** | **0.97** | **0.98** | **0.99** |

Given the high top-5 recall but relatively low top-1 recall for the crossencoder, an LLM was added for the final disease mapping(s). DeepSeek R1 (21) was used due to being open access and usable locally. A comparison was made to ChatGPT 5 (October 2025 version), with similar results, including a slightly higher F1 score (Table 3).

**Table 3:**
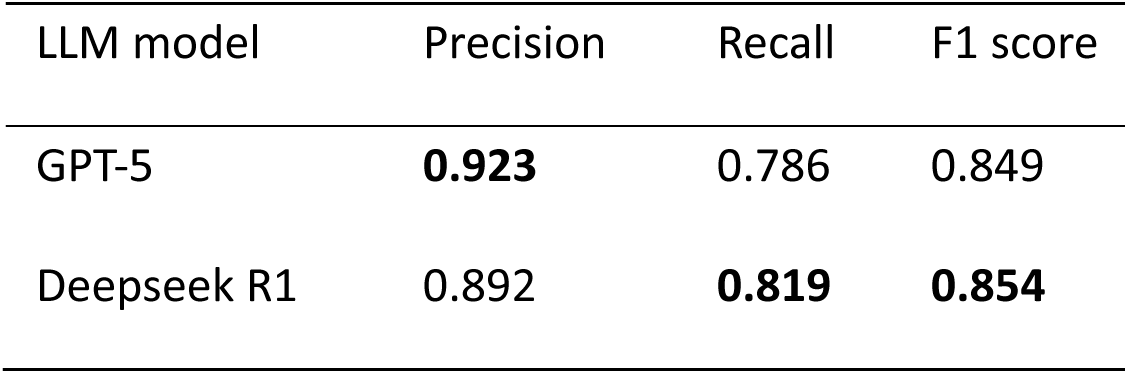
Ensemble disease mapping performance using Deepseek R1 (21) as final step in pipeline. Comparison made to ChatGPT 5 (31).

| LLM model | Precision | Recall | F1 score |
| --- | --- | --- | --- |
| GPT-5 | **0.923** | 0.786 | 0.849 |
| Deepseek R1 | 0.892 | **0.819** | **0.854** |

### Mapping DDG2P to PubMed

The pipeline described above was applied to all TIAB in PubMed. The resulting corpus of ∼69,000 papers, in theory, maps the publication landscape of all GDD in DDG2P. This data is available for download via the G2P website (https://www.ebi.ac.uk/gene2phenotype/). Results for each DDG2P entry can also be browsed on the website which is searchable on gene, phenotype or disease name. Currently, for disease mapped to large numbers of publications, only the latest 100 are displayed to improve usability. Analysis of this large-scale mapping found that diseases had more papers mapped using the automated method and the distribution of papers per disease was positively skewed (Figure 3), with the top five disorders by number of papers corresponding to well established conditions (Table 5). An example of differentiation between allelic disorders is shown in Table 4.

**Figure 3:**
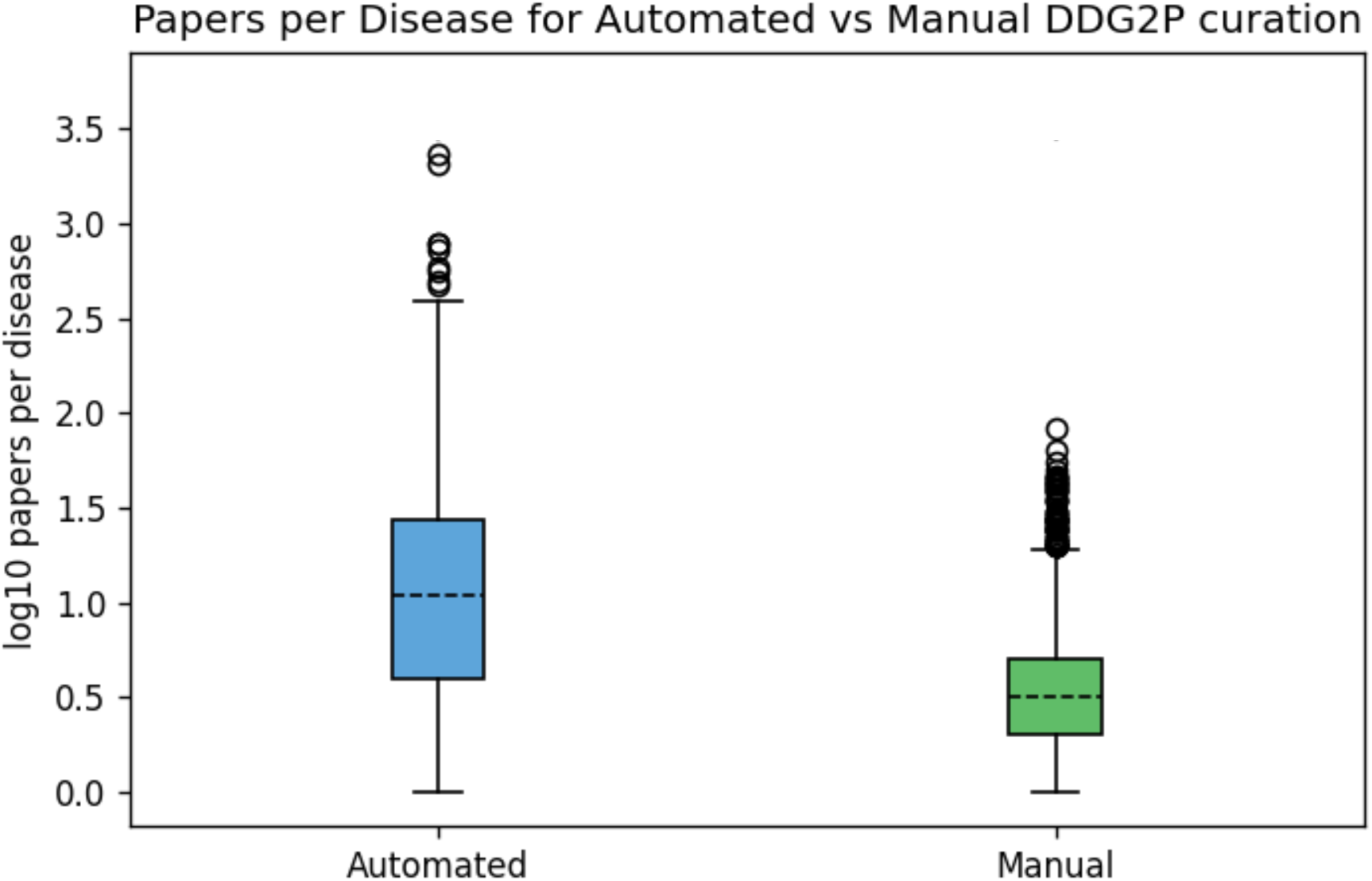
Comparison of papers per disease in existing manually curated DDG2P and with automated mapping. Log transform due to positive skew. Mann-Whitney U test p < 1e-300.

**Table 4:**
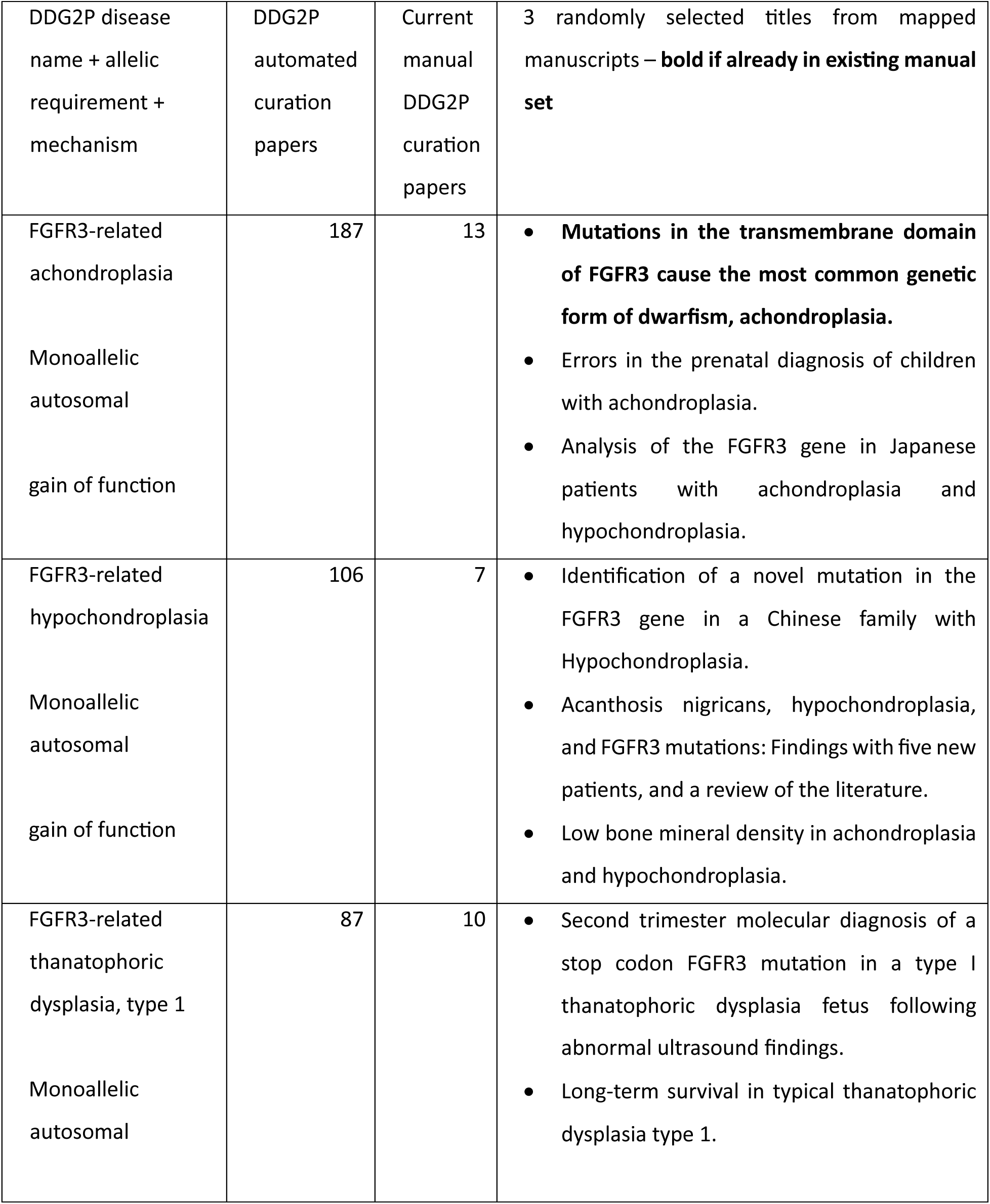
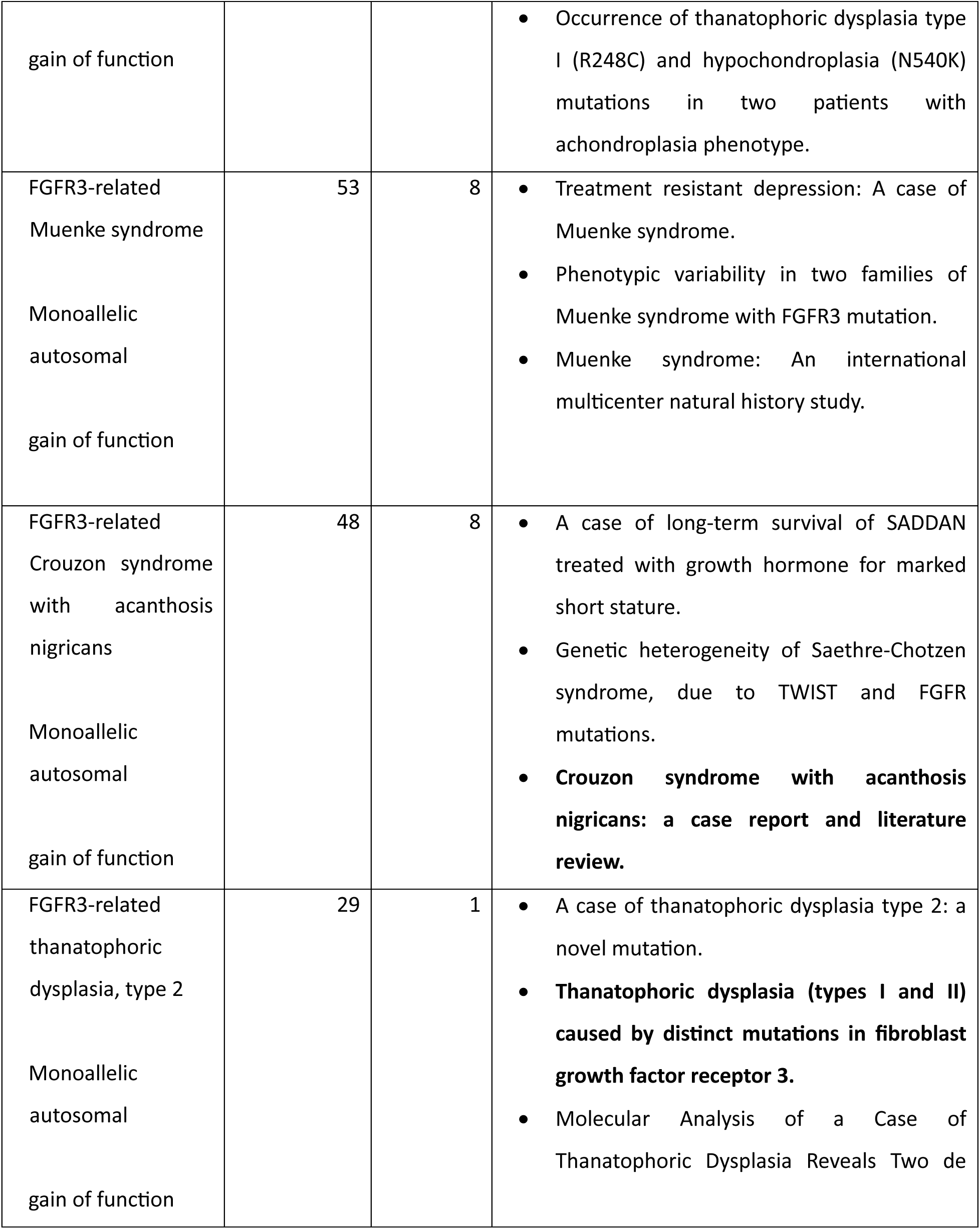
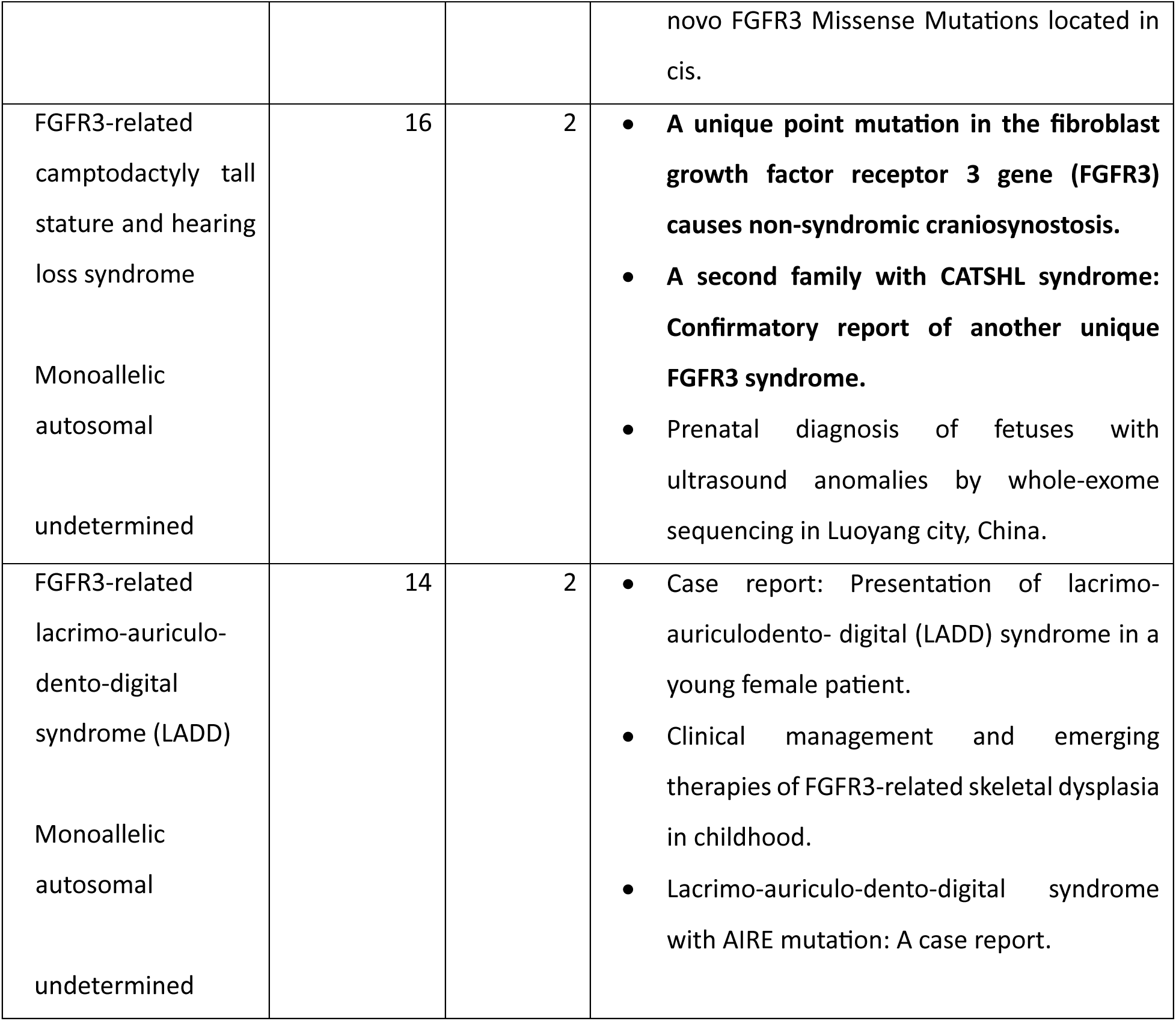
Example paper-disease mappings for allelic disorders in FGFR3. The challenge of mapping in these complex cases is illustrated e.g. ‘Genetic heterogeneity of Saethre-Chotzen syndrome, due to TWIST and FGFR mutations’ is mapped to ‘Crouzon syndrome with acanthosis nigricans’. Crouzon syndrome is mentioned in the abstract and the FGFR3 variant in the paper would now be described as causing Muenke syndrome.

**Table 5:**
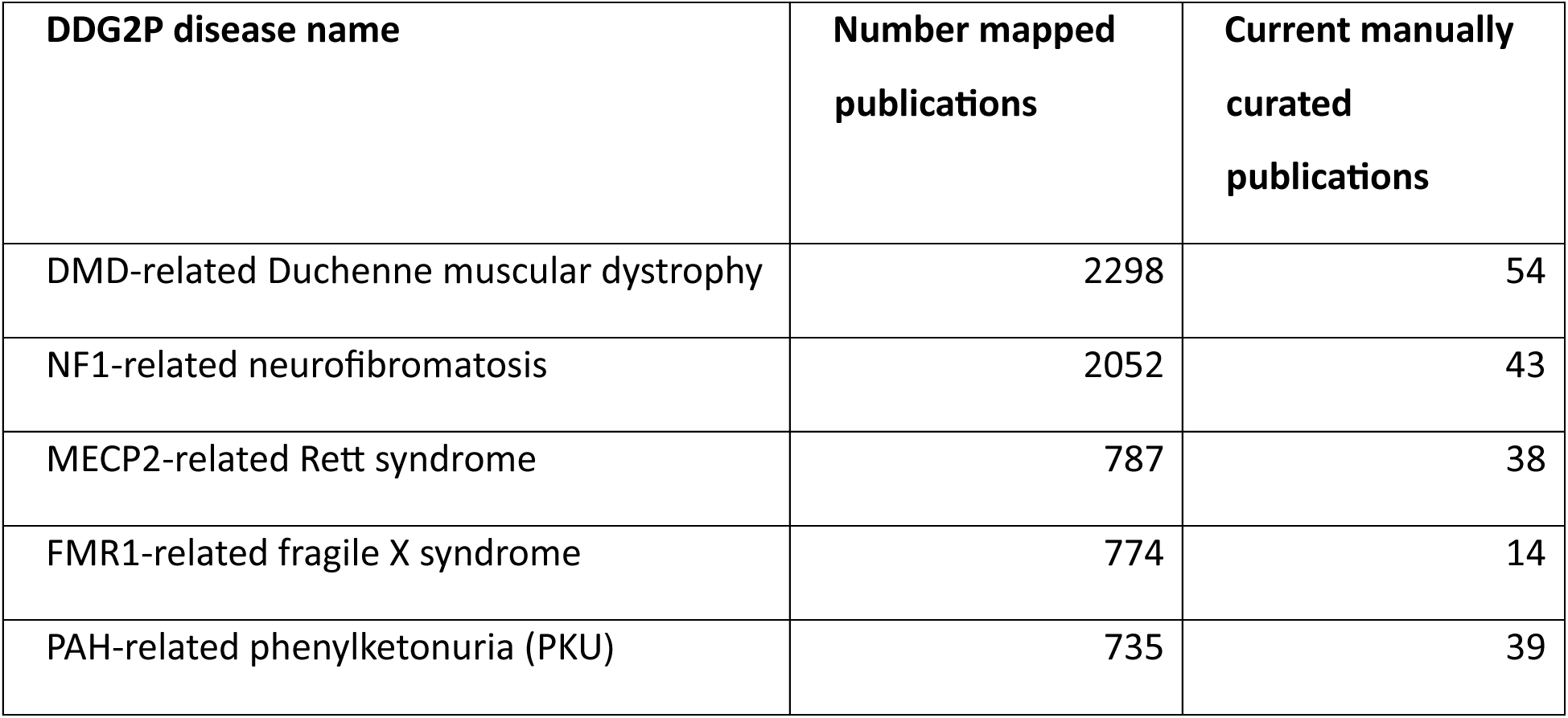
Top 5 diseases from auto curation by number of mapped publications. Note these are well-established, well-described disorders. These are outliers in the overall set where median papers per disease is 10 and mean 28.

A comparison was made with manually curated datasets to ensure the pipeline generalized to real-world applications (Table 6). Any manuscripts present in train/test data were excluded. Manuscripts already in DDG2P were reviewed (∼9000). Comparison was also made to external datasets. The Human Phenotype Ontology (HPO) provides a dataset of paper-disease-phenotype annotations, labelled by clinical experts (32). TIAB used for case level evidence in ClinGen gene-disease validity assessments were also analysed (7).

**Table 6:** Recall for auto-mapped literature disease pipeline vs existing manually curated data sources. Micro recall is overall proportion of PMIDs identified. Macro recall is mean per-disease recall.

| **Independent manually curated data source** | **Micro recall** | **Macro recall** |
| --- | --- | --- |
| Existing DDG2P | 0.70 | 0.71 |
| HPO Annotations | 0.68 | 0.72 |
| ClinGen | 0.68 | 0.66 |

## Discussion

Here, we have demonstrated the development and application of a system to address the challenge of identifying relevant manuscripts across the whole, highly heterogeneous, developmental disorders disease domain. Our approach allows for scalable automated search of the peer-reviewed literature, with mapping directly to curated diseases as defined in G2P, though it could be applied to other disease definitions with the same level of detail. We have applied our pipeline to the entirety of PubMed and made the resulting corpus freely available as part of the G2P database at https://www.ebi.ac.uk/gene2phenotype/. The manuscripts identified through our pipeline now form part of a curation workflow with manual review and annotation ongoing.

By combining a finetuned classifier (LitDD BERT), a crossencoder re-ranker (LitDD Crossencoder), and a constrained LLM step, the pipeline delivers high recall for retrieval from the entire biomedical literature, while preserving precision through multi-stage filtering and verification. On a held-out annotated test set, LitDD BERT achieved high discrimination with F1 approaching 0.9, indicating the classifier can parse out abstracts describing genetically determined disorders from millions of others in the literature. The crossencoder’s top-5 recall approached 1.0, supporting its use as a high-recall disease candidate generator. Because top-1 accuracy remained lower, we added an LLM to adjudicate among a small, high-quality candidate set. This led to a final mapping F1 of 0.85 with balanced precision and recall.

As with any approach built on foundation models, there is unavoidable overlap between the pretraining corpora and the evaluation literature. The large amount of data used to train LLMs is likely to include biomedical domain-specific text. Additionally, the use of baseline models which had already been finetuned for biomedical use likely results in training data overlap e.g. through use of the same abstracts. We minimized overfitting risk through stratified splits, conservative finetuning, and evaluation on unseen abstracts, but absolute performance may differ on corpora with different reporting styles. Generative models can hallucinate; we constrained the LLM to the top-5 crossencoder candidates, and verified the result to mitigate this risk. Here no finetuning was performed for the LLM step due to resource constraints. We plan in future to develop a finetuned domain-specific LLM which should increase precision and recall.

Our annotated training set was seeded with gene symbol[Title] queries, enriching for relevant case reports but biasing against abstracts that rely on disease names or syndromic labels. Disease names are heterogeneous and can be misaligned with molecular diagnosis. We intentionally avoided TIAB without explicit gene names/symbols for this reason.

Manual review of false positives and false negatives in the test set highlighted specific, recurring challenges. For false positives, these included copy-number variation (CNV) rather than single-nucleotide variants (SNV), often mentioning the correct gene, but not necessarily the correct phenotype. Also, somatic mutations were included in a few cases, from studies of common disease. Disease acronyms may be falsely identified as gene symbols. Finally, there were examples of correct gene but incorrect allelic disorder mappings (Table 4).

False negatives included mechanism mismatch, for example where the abstract supports haploinsufficiency or loss of function, but the relevant disorder was missense-only. There were also multi-map errors - TIAB describing a single allelic entity mapped to multiple diseases and vice versa. For X linked conditions, TIAB describing males were mapped to female-only conditions another example of allelic disorder error.

Additionally, 100 TIAB randomly chosen from the final mapped set were reviewed. Similar themes for a minority of incorrect mappings emerged, including CNV rather than SNV, multi-disease map mismatch. Examples of allelic adult-onset conditions being mapped to DDG2P paediatric conditions were also observed.

These themes underscore that the main source of error is disambiguation among allelic disorders and mechanism-specific disease definitions. Disease naming is inconsistent across the literature and remains a source of debate (16,17). Lumping and splitting allelic disorders is a known challenge in genetic disease (18). Also, mechanism is not always explicitly mentioned in TIAB. Therefore, it is not surprising this is a challenge for our pipeline. We plan to develop case-level extraction to mitigate many of these errors by explicitly capturing variant class, zygosity, mechanism, sex, age-of-onset, and ascertainment context (likely using full text), and then aligning these against the LGMDE definition. This will allow further standardization through generation of data compatible with community standards such as Phenopackets (33).

Comparison of our automated curation approach to others is confounded by their method of mapping to genes only, rather than disease. AMELIE used a logistic regression classifier to identify more than 500,000 articles from 29 million PubMed abstracts, mapped to gene (38). This is far more than our set and likely reflects, in part, inclusion of non-human and mechanistic manuscripts. EvAgg uses a pure LLM approach to search by gene for relevant papers (13). We attempted to use EvAgg on our labelled set but ran into unresolvable NCBI API errors. The gold standard of paper identification, however, remains manual curation. In evaluation against real-world unseen curated sets (7,8,32) the pipeline recovered the majority of known PMIDs (micro and macro recall around 0.70), supporting generalizability across heterogeneous disease mechanisms, inheritance patterns, and curation formats. There are likely multiple reasons for the missed 30% of papers. For example, 13 papers in DDG2P were referenced by five or more (max 48) disease entries. This reflects inclusion of large cohort studies which do not reliably contain case-level data and which the pipeline here was not designed to identify. We also use TIAB only, and do not make use of full-text or supplementary data. TIAB do not always contain information pertaining to individual diseases and expansion to available full text in future is likely to increase recall.

Applying the pipeline to all of PubMed produced a corpus of approximately 69,000 manuscripts mapped to DDG2P diseases. The overview map (Figure 4) gives an intuition as to one of the potential uses of this data - mapping individuals to a disease cluster via phenotype should narrow the genomic search space and guide follow-up testing.

**Figure 4:**
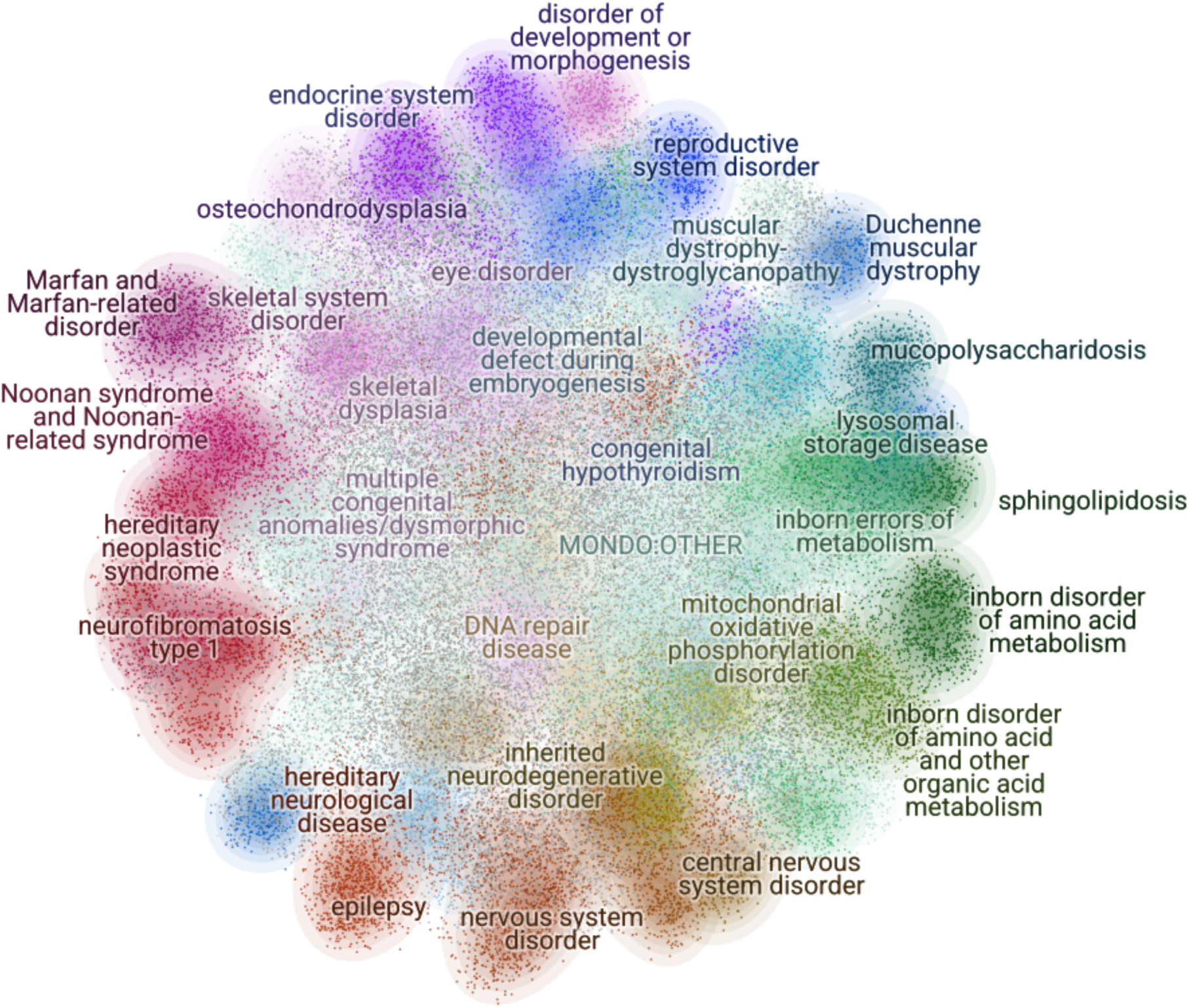
DDG2P PubMed map. ∼69,000 TIAB identified using classification and disease map pipeline. Text embeddings created using BioClinical ModernBERT (34). Mondo-G2P mappings used to collapse cluster labels to lowest shared common ancestor (35). NB MONDO:OTHER label is for TIAB with only root as shared ancestor. Clustering was using HDBSCAN and 2D map with UMAP (36,37).

This collection of publications is, more comprehensive than those in the existing manually curated databases (6–8), as these explicitly do not attempt to identify all publications linked to a given disease, in contrast to our work. We show that automated curation gives more manuscripts per disease than the manual approach for G2P (Figure 3). This means the domain-wide mappings should provide a solid foundation for biomedical knowledge graphs. We intend to run phenotype extraction on this set, with the aim of both improving the results of existing disease predictors such as Exomiser (39) and developing novel algorithms for this purpose. Because the process is automated and modular, it can be re-run regularly as PubMed and DDG2P are updated, supporting continuous literature surveillance for biocurators and clinicians.

In summary, the presented pipeline delivers scalable, automated literature identification and disease mapping for developmental disorders. This is already augmenting the G2P curation process. The principal remaining challenge is precise disambiguation among allelic disorders and including edge cases such as mosaicism, complex structural variation and overlapping phenotypes. By integrating case-level extraction, we anticipate further gains in coverage, precision, and clinical utility.

## Data Availability

All PMID-disease mappings for DDG2P are available to download from the G2P website https://www.ebi.ac.uk/gene2phenotype/.
Code to replicate this study, as well as the annotated corpus and final disease mappings is available at https://github.com/biomedicalinformaticsgroup/ LitDD_mining.
The LitDD BERT and LitDD crossencoder models are available from huggingface: https://huggingface.co/tmy100000001/LitDD_BERT/
https://huggingface.co/tmy100000001/LitDD_crossencoder

https://github.com/biomedicalinformaticsgroup/LitDD_mining

https://huggingface.co/tmy100000001/LitDD_BERT/

https://huggingface.co/tmy100000001/LitDD_crossencoder

## Acknowledgments

This research was funded in whole or in part by the Wellcome Trust [226083/Z/22/Z] for PARADIGM (Primary Annotated Resources to Advance Discovery In Genomic Medicine). For the purpose of open access, the author has applied a CC-BY public copyright licence to any author accepted manuscript version arising from this submission. HVF was supported by the NIHR Cambridge Biomedical Research Centre (NIHR203312) and CFW was supported by the NIHR Exeter Biomedical Research Centre The views expressed are those of the authors and not necessarily those of the NIHR or the Department of Health and Social Care.

## Web resources

All PMID-disease mappings for DDG2P are available to download from the G2P website https://www.ebi.ac.uk/gene2phenotype/.

## Data and code availability

Code to replicate this study, as well as the annotated corpus and final disease mappings is available at https://github.com/biomedicalinformaticsgroup/ LitDD_mining.

The LitDD BERT and LitDD crossencoder models are available from huggingface: https://huggingface.co/tmy100000001/LitDD_BERT/ https://huggingface.co/tmy100000001/LitDD_crossencoder

## References

1. The 1000 Genomes Project Consortium, Corresponding authors, Auton A, Abecasis GR, Steering committee, Altshuler DM, et al. A global reference for human genetic variation. Nature. 2015 Oct 1;526(7571):68–74.

2. Thormann A, Halachev M, McLaren W, Moore DJ, Svinti V, Campbell A, et al. Flexible and scalable diagnostic filtering of genomic variants using G2P with Ensembl VEP. Nat Commun. 2019 May 1;10(1):2373.

3. Wright CF, Campbell P, Eberhardt RY, Aitken S, Perrett D, Brent S, et al. Genomic Diagnosis of Rare Pediatric Disease in the United Kingdom and Ireland. N Engl J Med. 2023 Apr 27;388(17):1559–71.

4. Wilczewski CM, Obasohan J, Paschall JE, Zhang S, Singh S, Maxwell GL, et al. Genotype first: Clinical genomics research through a reverse phenotyping approach. Am J Hum Genet. 2023 Jan;110(1):3–12.

5. Durkie M, Cassidy EJ, Berry I, Owens M, Turnbull C, Taylor RW, et al. ACGS Best Practice Guidelines for Variant Classification in Rare Disease 2024 [Internet]. 2024 [cited 2025 Aug 18]. Available from: https://www.acgs.uk.com/media/12533/_media_12533_uk-practice-guidelines-for-variant-classification-v12-2024.pdf

6. Amberger JS, Bocchini CA, Scott AF, Hamosh A. OMIM.org: leveraging knowledge across phenotype–gene relationships. Nucleic Acids Res. 2019 Jan 8;47(D1):D1038–43.

7. Andersen EF, Azzariti DR, Babb L, Berg JS, Biesecker LG, Bly Z, et al. The Clinical Genome Resource (ClinGen): Advancing genomic knowledge through global curation. Genet Med. 2025 Jan;27(1):101228.

8. Yates TM, Ansari M, Thompson L, Hunt SE, Uhalte EC, Hobson RJ, et al. Curating genomic disease-gene relationships with Gene2Phenotype (G2P). Genome Med. 2024 Nov 6;16(1):127.

9. DiStefano MT, Goehringer S, Babb L, Alkuraya FS, Amberger J, Amin M, et al. The Gene Curation Coalition: A global effort to harmonize gene-disease evidence resources [Internet]. Genetic and Genomic Medicine; 2022 Jan [cited 2022 Jan 5]. Available from: http://medrxiv.org/lookup/doi/10.1101/2022.01.03.21268593

10. Sayers EW, Bolton EE, Brister JR, Canese K, Chan J, Comeau DC, et al. Database resources of the national center for biotechnology information. Nucleic Acids Res. 2021 Dec 1;gkab1112.

11. Qin X, Liu J, Wang Y, Liu Y, Deng K, Ma Y, et al. Natural language processing was effective in assisting rapid title and abstract screening when updating systematic reviews. J Clin Epidemiol. 2021 May;133:121–9.

12. Masoumi S, Amirkhani H, Sadeghian N, Shahraz S. Natural language processing (NLP) to facilitate abstract review in medical research: the application of BioBERT to exploring the 20-year use of NLP in medical research. Syst Rev. 2024 Apr 15;13(1):107.

13. Twede H, Conard AM, Pais L, Bryen S, O’Heir E, Smith G, et al. Evidence Aggregator: AI reasoning applied to rare disease diagnostics [Internet]. 2025 [cited 2025 Mar 18]. Available from: http://biorxiv.org/lookup/doi/10.1101/2025.03.10.642480

14. Pilehvar MT, Bernard A, Smedley D, Collier N. PheneBank: a literature-based database of phenotypes. Wren J, editor. Bioinformatics. 2022 Jan 27;38(4):1179–80.

15. Wei CH, Allot A, Lai PT, Leaman R, Tian S, Luo L, et al. PubTator 3.0: an AI-powered literature resource for unlocking biomedical knowledge. Nucleic Acids Res. 2024 July 5;52(W1):W540– 6.

16. Biesecker LG, Adam MP, Alkuraya FS, Amemiya AR, Bamshad MJ, Beck AE, et al. A dyadic approach to the delineation of diagnostic entities in clinical genomics. Am J Hum Genet. 2021 Jan 7;108(1):8–15.

17. Hamosh A, Amberger JS, Bocchini CA, Bodurtha J, Bult CJ, Chute CG, et al. Response to Biesecker et al. Am J Hum Genet. 2021 Sept 2;108(9):1807–8.

18. Thaxton C, Goldstein J, DiStefano M, Wallace K, Witmer PD, Haendel MA, et al. Lumping versus splitting: How to approach defining a disease to enable accurate genomic curation. Cell Genomics. 2022 May;2(5):100131.

19. Warner B, Chaffin A, Clavié B, Weller O, Hallström O, Taghadouini S, et al. Smarter, Better, Faster, Longer: A Modern Bidirectional Encoder for Fast, Memory Efficient, and Long Context Finetuning and Inference [Internet]. arXiv; 2024 [cited 2025 Oct 20]. Available from: http://arxiv.org/abs/2412.13663

20. Jin Q, Kim W, Chen Q, Comeau DC, Yeganova L, Wilbur WJ, et al. MedCPT: Contrastive Pre-trained Transformers with large-scale PubMed search logs for zero-shot biomedical information retrieval. Wren J, editor. Bioinformatics. 2023 Nov 1;39(11):btad651.

21. DeepSeek-AI, Guo D, Yang D, Zhang H, Song J, Zhang R, et al. DeepSeek-R1: Incentivizing Reasoning Capability in LLMs via Reinforcement Learning [Internet]. arXiv; 2025 [cited 2025 Aug 28]. Available from: https://arxiv.org/abs/2501.12948

22. Seal RL, Braschi B, Gray K, Jones TEM, Tweedie S, Haim-Vilmovsky L, et al. Genenames.org: the HGNC resources in 2023. Nucleic Acids Res. 2023 Jan 6;51(D1):D1003–9.

23. Wolf T, Debut L, Sanh V, Chaumond J, Delangue C, Moi A, et al. HuggingFace’s Transformers: State-of-the-art Natural Language Processing [Internet]. arXiv; 2019 [cited 2025 Jan 10]. Available from: https://arxiv.org/abs/1910.03771

24. Liaw R, Liang E, Nishihara R, Moritz P, Gonzalez JE, Stoica I. Tune: A Research Platform for Distributed Model Selection and Training [Internet]. arXiv; 2018 [cited 2025 Jan 10]. Available from: https://arxiv.org/abs/1807.05118

25. Balachandran, Abhinand. MedEmbed: Medical-Focused Embedding Models [Internet]. 2024. Available from: https://github.com/abhinand5/MedEmbed

26. Moreira G de SP, Osmulski R, Xu M, Ak R, Schifferer B, Oldridge E. NV-Retriever: Improving text embedding models with effective hard-negative mining [Internet]. arXiv; 2024 [cited 2025 Sept 1]. Available from: https://arxiv.org/abs/2407.15831

27. Lee J, Yoon W, Kim S, Kim D, Kim S, So CH, et al. BioBERT: a pre-trained biomedical language representation model for biomedical text mining. Wren J, editor. Bioinformatics. 2019 Sept 10;btz682.

28. Gu Y, Tinn R, Cheng H, Lucas M, Usuyama N, Liu X, et al. Domain-Specific Language Model Pretraining for Biomedical Natural Language Processing. ACM Trans Comput Healthc. 2022 Jan 31;3(1):1–23.

29. Reimers N, Gurevych I. Sentence-BERT: Sentence Embeddings using Siamese BERT-Networks [Internet]. arXiv; 2019 [cited 2025 Oct 20]. Available from: http://arxiv.org/abs/1908.10084

30. Kyei-Mensah, Bernard. medimaven-reranker-bge-cross-encoder [Internet]. 2025. Available from: https://huggingface.co/dranreb1660/medimaven-reranker-bge-cross-encoder

31. OpenAI. ChatGPT 5 (October 2025 version) [Internet]. 2025. Available from: https://chat.openai.com/chat

32. Köhler S, Gargano M, Matentzoglu N, Carmody LC, Lewis-Smith D, Vasilevsky NA, et al. The Human Phenotype Ontology in 2021. Nucleic Acids Res. 2021 Jan 8;49(D1):D1207–17.

33. Jacobsen JOB, Baudis M, Baynam GS, Beckmann JS, Beltran S, Buske OJ, et al. The GA4GH Phenopacket schema defines a computable representation of clinical data. Nat Biotechnol. 2022 June;40(6):817–20.

34. NeuML. BioClinical ModernBERT [Internet]. 2025. Available from: https://huggingface.co/NeuML/bioclinical-modernbert-base-embeddings

35. Shefchek KA, Harris NL, Gargano M, Matentzoglu N, Unni D, Brush M, et al. The Monarch Initiative in 2019: an integrative data and analytic platform connecting phenotypes to genotypes across species. Nucleic Acids Res. 2020 Jan 8;48(D1):D704–15.

36. Bot DM, Peeters J, Liesenborgs J, Aerts J. FLASC: a flare-sensitive clustering algorithm. PeerJ Comput Sci. 2025 Apr 18;11:e2792.

37. Sainburg T, McInnes L, Gentner TQ. Parametric UMAP embeddings for representation and semi-supervised learning [Internet]. arXiv; 2020 [cited 2025 Oct 31]. Available from: https://arxiv.org/abs/2009.12981

38. Birgmeier J, Haeussler M, Deisseroth CA, Steinberg EH, Jagadeesh KA, Ratner AJ, et al. AMELIE speeds Mendelian diagnosis by matching patient phenotype and genotype to primary literature. Sci Transl Med. 2020 May 20;12(544):eaau9113.

39. Smedley D, Jacobsen JOB, Jäger M, Köhler S, Holtgrewe M, Schubach M, et al. Next-generation diagnostics and disease-gene discovery with the Exomiser. Nat Protoc. 2015 Dec;10(12):2004–15.

